# Network structure induced bias in estimates of intrinsic generation times

**DOI:** 10.1101/2025.05.15.25327595

**Authors:** Pratyush K. Kollepara, Chiara Poletto, Joel C. Miller

## Abstract

The reproduction number is a critical measure of the severity of an infectious disease epidemic. The generation interval, defined as the time taken by an infector to create another infection from its time of infection, is crucial for estimating the basic reproduction number. However, the generation intervals observed, ‘realised generation intervals’, change over time depending on the dynamics of the epidemic. These time varying distributions are well understood for homogeneous populations, and can be used to infer the intrinsic generation interval distribution. For heterogeneous populations, the state-of-the-art method relies on the use of expensive network based or agent based simulations. We simplify this process by writing exact expressions for simple structured populations. We then use these expressions to demonstrate some previously unexplored biases in the estimation of the intrinsic generation times from the observed generation times, caused by the network structure assumed in the model and particularly the temporal structure of contacts.

## 1 Introduction

Generation time, the time a parent takes to produce an offspring since its birth, is an important quantity of interest in ecology. The concept has been used extensively in epidemiology, where it refers to the time taken for a newly infected individual to infect someone else. Knowing this interval distribution is necessary for planning contact tracing, quarantine, and isolation in case of an outbreak [13]. It is also necessary to estimate ℛ_0_, which quantifies the epidemic transmission potential, from the aggregated incidence of infections using the Wallinga-Lipsitch equation [15]. However, measuring this quantity and correctly interpreting it for outbreak analysis is not straightforward [17].

The Wallinga-Lipstich equation requires the “intrinsic generation interval”, which is defined as the time distribution of infectious contacts made by a primary case, or, equivalently, the time distribution of generated infections when the population is fully susceptible and mixes randomly [17]. The observed or realised generation intervals, obtained by contact tracing data, are different from the intrinsic ones, because of dynamical and population mixing effects, and also because of how generation intervals are defined and aggregated from available data. For instance, The forward generation interval distribution refers to the distribution of times it takes for a group of individuals infected at a given time to transmit the infection to others. On the other hand, the backward generation interval distribution takes the point of view of a cohort of infectees, infected at a given time, and is defined as the distribution of times to infect them [17].

The intrinsic generation interval, determined solely by the infector’s infectiousness, remains constant throughout an outbreak. In contrast, forward and backward realized generation times vary. The functional relationship between intrinsic and realised generation time was mathematically studied for a randomly mixing population by the Champredon and Dushoff [17] within the renewal equation framework. The backward generation time increases monotonically, underestimating the intrinsic generation interval early on and overestimating it later. This bias arises from the incidence dynamics. During the growth phase, recent infections dominate the pool of potential infectors, skewing generation times shorter; during the decline, older infections dominate, lengthening the generation intervals. On the other hand, the forward-looking formulation of generation time avoids this dynamic bias. Still, its mean value shrinks right before the peak [17, 16, 12]. One reason for this contraction is the high rate at which susceptibles are being depleted near the peak. An infector may generate multiple transmissions throughout their infectious period and, if the number of susceptibles remains the same, then the transmission events are all equally likely to contribute to the generation intervals [17]. Conversely, if the availability of susceptibles shrinks significantly during the infectious period the later transmissions are less likely to lead to an infection and longer transmission intervals are less likely to be sampled. An alternative mechanism to understand the generation time contraction is the competition between infectors, racing to infect someone [16, 11]. When many infectors compete to infect the same susceptible individual, only the transmission by the fastest infector counts as an infection event and contributes to the generation time statistics, causing the generation interval to contract. The study of these dynamical mechanisms is essential to design techniques to fit the intrinsic generation time from contact tracing data [5, 14, 6].

Although most studies rely on the homogenous mixing assumption, contacts within households, schools, and workplaces are recurrent, heterogeneous, and spatially clustered [19, 3, 9, 10, 7, 8, 1, 2, 19, 4]. Previous studies investigating clustering and recurrence showed that these enhance the competition between infectors and cause a stronger reduction of generation intervals — more than predicted by homogeneous-mixing models — which persists throughout the outbreak [19, 11]. Despite these findings, the effect of the contact network on the generation time dynamics remains under-studied, especially from an analytical perspective. Here, we use the edge-based compartmental modelling framework [18] to derive exact equations for realized generation intervals. The framework allows us to systematically address the role of contacts’ heterogeneity - by comparing a homogeneous and a heterogeneous network - and recurrence - by comparing the cases in which contacts’ identities continuously change or are fixed. Our theoretical framework, supported by simulations, provides insights into the dynamics of generation intervals and highlights differences between network topologies. Most importantly it points to potential sources of bias, that were left unexplored so far.

## 2 Methods

This section gives a brief overview of the Edge Based Compartmental Modelling (EBCM) framework [18] that is required for understanding the results in the next section. The EBCM framework can be used to solve the SIR model on configuration model networks. These networks are specified by a distribution of the degrees of nodes in the network but are limited to a tree-like structure (do not allow clustering or higher-order structures).

Real-world networks have a temporal nature: some contacts of an individual are fleeting (fast changing) while others are static (long-lasting). The models used here will consider versions of configuration model networks for each type of contact. The first one is known as the annealed or mean field configuration model where even though the number of contacts of an individual remains constant, the edges in the network rewire to new nodes constantly. The other is known as the quenched or static configuration model where the edges are static. In addition to these assumptions about the network structure, we also assume that both transmission and recovery from infection are described by a Poisson process. Detailed derivations of our results are presented in the Supplementary Methods section, and the results can be generalised to non-Poisson processes.

For both annealed and quenched networks, a standard compartmental modelling approach would divide the population up into sub-groups of homogeneous degree (number of contacts), resulting in at least as many differential equations as the number of sub-groups. The EBCM framework reduces such a large system of differential equations into two ordinary differential equations which can be solved numerically with ease.

The EBCM framework makes use of probability-generating functions

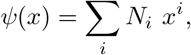

where *N*_*i*_ refers to the proportion or probability of nodes of degree class *i*. A variable *θ* is defined to be the probability that a randomly chosen partnership of a node has not transmitted to it yet. If a node of degree *k* is susceptible, then none of its *k* partners have transmitted to it. The probability for the node to be susceptible (given its degree *k*) is *θ*^*k*^. The prevalence of susceptibles in the population can be computed using

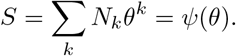

### 2.1 Annealed network

Homogeneous compartmental models use macroscopic variables such as the proportion of the population that is infected, susceptible, recovered etc. The probability a random interaction with another individual is with an infected individual is assumed to equal the proportion of the population that is infected. In a more general case where the number of partners differs among individuals, the probability a random contact is with an infected individual is the prevalence of half-edges in the population that are attached to an infected node. The variable *I*(*t*) (the proportion of the population that is infected) is generalised to a variable *π*_*I*_ (*t*) which is the proportion of half-edges that connect to infected nodes. Similarly, we generalize *π*_*S*_(*t*) and *π*_*R*_(*t*) which are the prevalence of half-edges with susceptible and recovered nodes respectively. We also define a new variable *π*_*i*_(*t*), which is the incidence of infected stubs, i.e. the instanteous rate at which half-edges with infected nodes are created by new infections.

We find that the probability a random half-edge connects to a susceptible node is

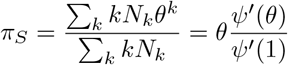

Knowing *π*_*S*_ requires *θ*(*t*). At the start of the epidemic, the initially susceptible nodes have not received any transmissions. So, using the initial condition *θ*(0) ≈ 1, the following differential equation (derived in ref [18]) can be solved to find *θ*(*t*):

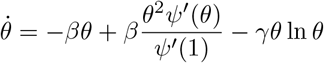

### 2.2 Quenched network

In the quenched case, the variable *S*(*t*) is generalised to a variable *ϕ*_*S*_(*t*) which is the probability that the partner of a randomly selected node is susceptible (under the assumption that the randomly selected node has not transmitted to its partner).

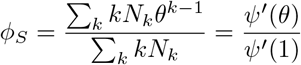

In this case, using a similar initial condition for *θ* as before, the following differential equation [18] is solved:

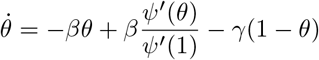

## 3 Results

### 3.1 Annealed network

In a homogeneous population, the forward generation interval distribution can be calculated using the intrinsic generation interval distribution, *g*(*τ*), and the proportion or number of susceptible people, *S*(*t*), with time [17].

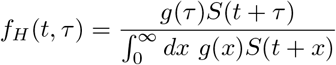

Here, *S*(*t*) gives the probability for a random individual to come in contact with a susceptible individual at time *t*. In an annealed heterogeneous network this is no longer *S*(*t*) and depends on the connectivity. We thus need to replace it with *π*_*S*_(*t*). Therefore, the forward generation interval distribution for an annealed configuration model network is

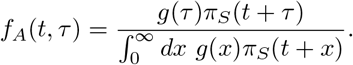

Turning our attention to the backward generation interval, Champredon and Dushoff’s equation for the homogeneous case makes use of the incidence of new infections as a function of time [17]:

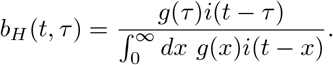

If we use the average incidence of stubs, instead of the incidence of infections, we obtain an exact expression for the backward generation interval distribution for an annealed heterogeneous network.

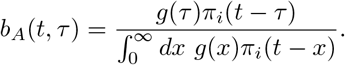

A general characteristic of the backward generation intervals is that they are stable in time when the incidence grows exponentially [17]. This remains valid in the heterogeneous network case, as can be seen by plugging in *π*_*i*_(*t*) = *π*_*i*_(0)*e*^*λt*^ into the above formula, leading to

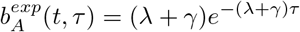

Similar to the forward generation interval, we define an effective reproduction number called the forward reproduction number, ℛ_*f*_. It represents the expected number of infections created by an average infector who became infected at time *t* over its entire infectious period. For an annealed network,

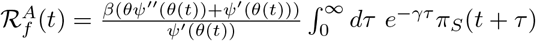

If a reproduction number is calculated empirically through observation of transmission chains, we would expect it to match with the forward reproduction number.

### 3.2 Quenched network

In an annealed network, a node continuously breaks its edges and re-connects to new edges. So, if a node infects another node, it does not compete with itself when transmitting again. Conversely, in a quenched network, a node can compete with itself — after the first transmission all later transmissions to the same neighbour have no effect. In the equations for the realised generation time we thus need to replace the intrinsic generation time *g*(*τ*) with 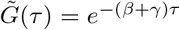. In addition, the probability that a stub points to a susceptible node – which was *π*_*S*_(*t*) in the annealed case – is now given by *ϕ*_*S*_(*t*). The modified version of the equation in [17] then becomes

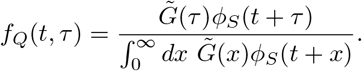

In the backward case, the incidence of infections, *i*(*t*) is replaced with the average probability of incidence among the neighbours of a node, *ϕ*_*i*_(*t*):

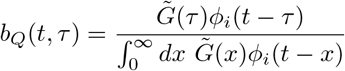

From this equation, we obtain that the backward generation intervals are stable when the epidemic grows exponentially, similar to the annealed case. The distribution in this regime is

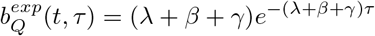

Finally, the forward reproduction number can be derived for quenched network as follows

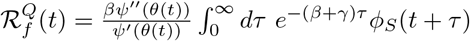

### 3.3 Impact of network properties on the realised generation time

We validate the exact results using event-based simulations [21]. Figures 1 and 2 show the comparison between analytical solutions and simulations for the annealed and quenched case, respectively. In each figure, we plot the epidemic profile, the forward and backward generation times and the forward reproduction number. The figures show the comparison for two different topologies that we call TMD and TPL70. TMD refers to a tri-modal degree distribution and TPL70 refers to a truncated power law degree distribution (See Figure captions more details). Both networks are heterogeneous, but the TPL70 network is a case of extreme heterogeneity. The forward generation intervals in the annealed case show a contraction qualitatively similar to what is predicted by the homogenous mixing equations [17]. This is, however, less evident when the network is extremely heterogeneous. In the quenched case forward generation times shrink substantially due to the intensified competition among infectors. While forward generation time varies substantially from one network to another, backwards generation time is quite robust to the network structure.

**Figure 1:**
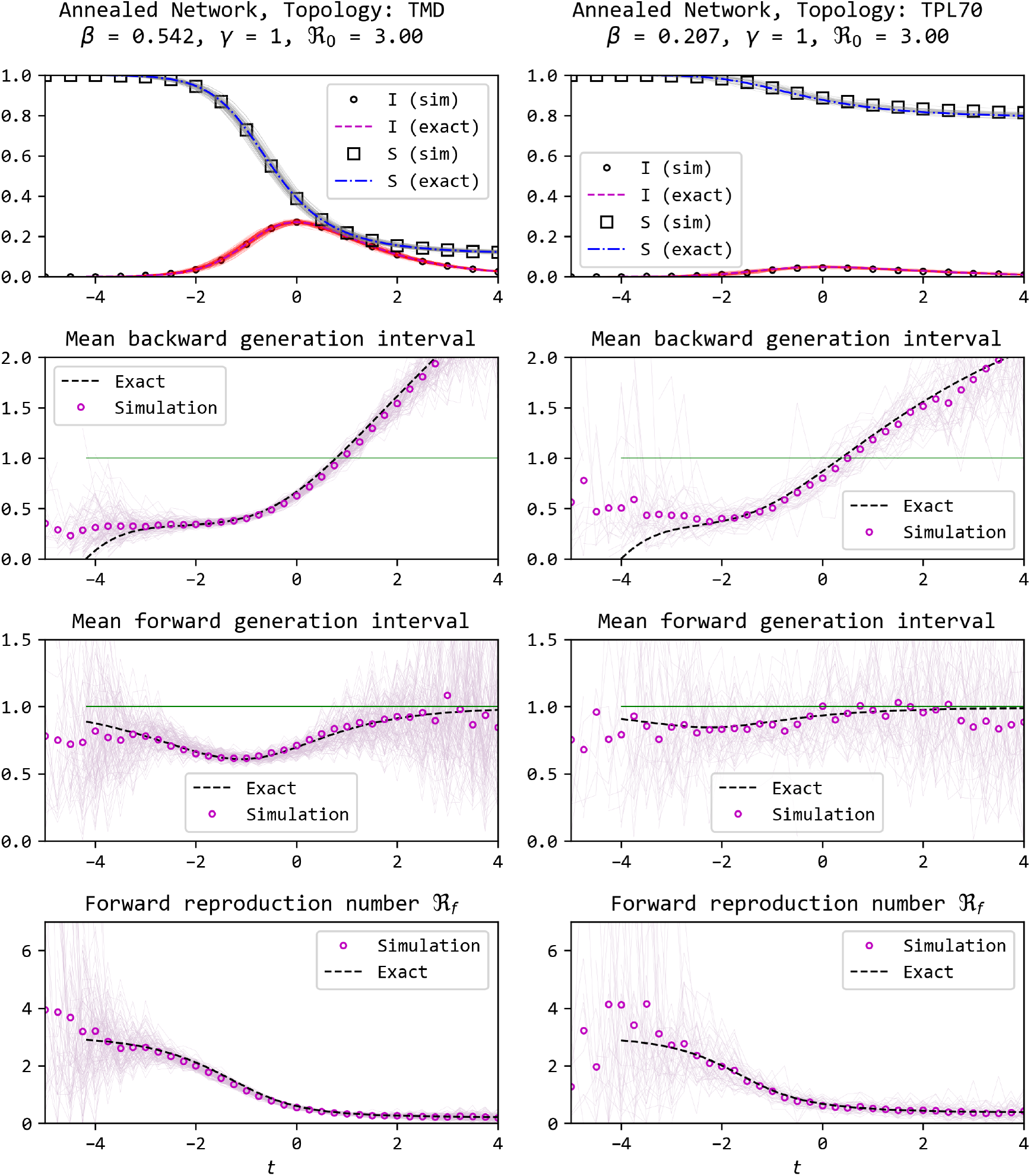
Comparison of epidemics and their realised generation intervals using an event-based simulation method and the exact solution for annealed networks with two types of heterogeneous degree distributions (Left: the degree of a node can be 3, 5, or 7 with equal probabilities, Right: degree distribution is a truncated power law with maximum degree of 70 and exponent of −2). We find a good match between theory and simulations for the realised generation intervals and the forward reproduction number. The time-series are centred such that the prevalence peaks at time zero. The thin lines show trajectories from each of the 100 simulations, while the plot with markers shows the average. At the start and towards the end of the epidemic, significant noise is observed and the simulation deviates from the theory.

**Figure 2:**
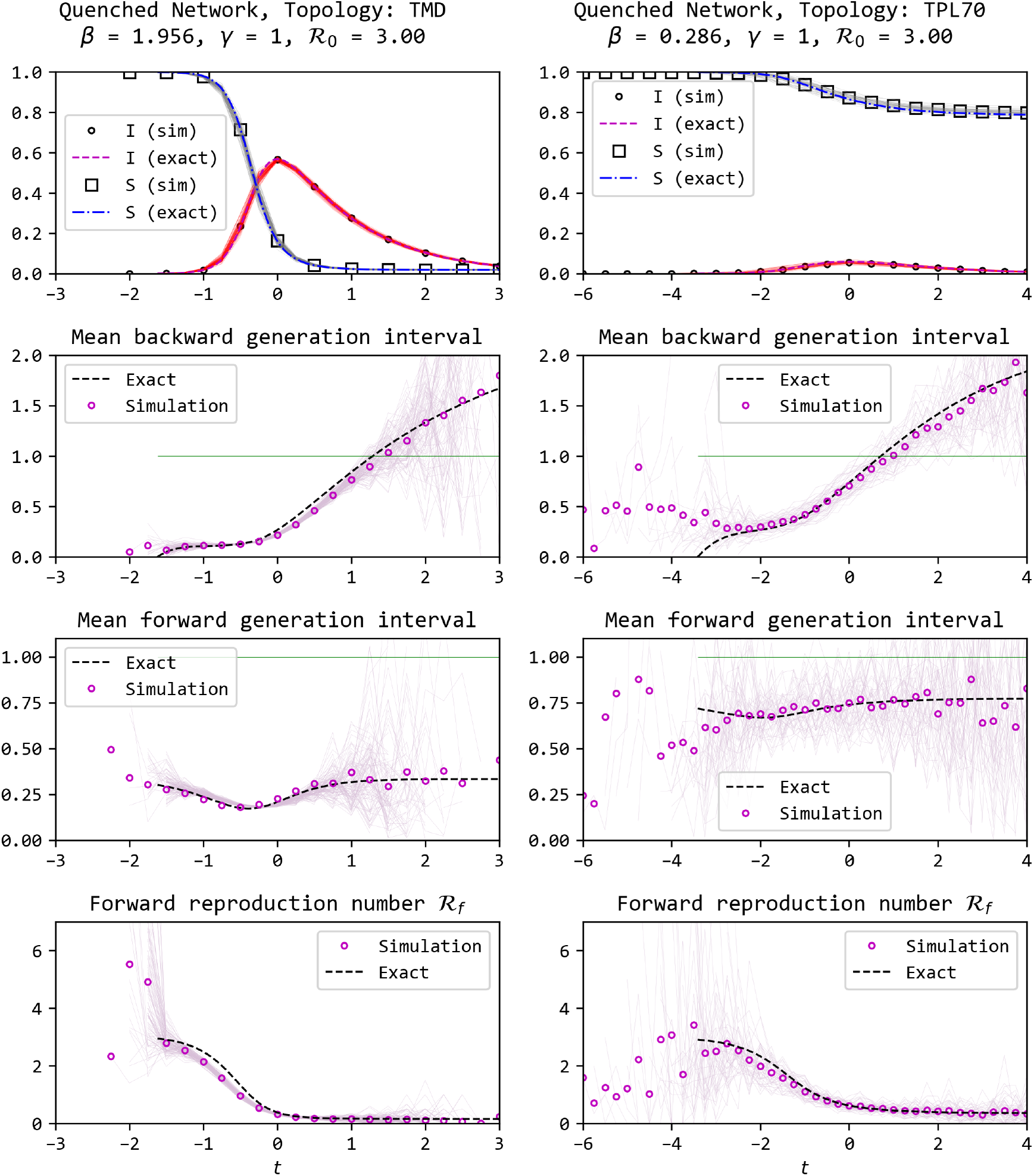
Comparison of epidemics and their realised generation intervals using an event-based simulation method and the exact solution for quenched networks with two types of heterogeneous degree distributions (Left: the degree of a node can be 3, 5, or 7 with equal probabilities, Right: degree distribution is a truncated power law with maximum degree of 70 and exponent of −2). We find a good match between theory and simulations for the realised generation intervals and the forward reproduction number. The time-series are centred such that the prevalence peaks at time zero. The thin lines show trajectories from each of the 100 simulations, while the plot with markers shows the average. At the start and towards the end of the epidemic, significant noise is observed and the simulation deviates from the theory for the truncated power law distribution.

**Figure 3:**
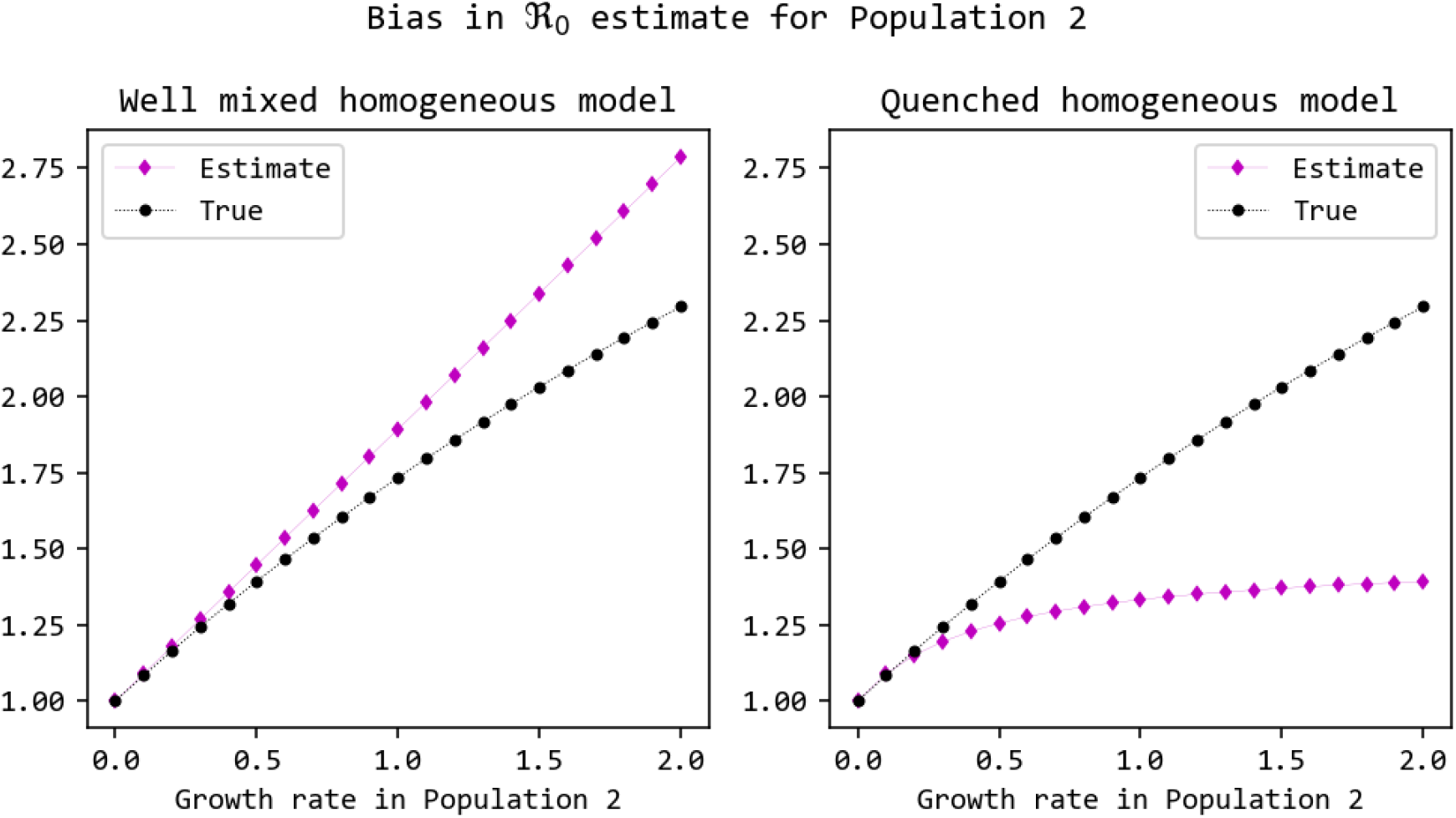
Comparison between the estimated and true value of the basic reproduction number in a heterogeneous quenched network when different homogeneous models are used and the intrinsic generation interval is estimated from a different population (see 3.4 for a detailed explanation).

We can further analyse the impact of the network features by numerically solving the derived analytical expressions for different scenarios. This is significantly inexpensive compared to stochastic numerical simulations. We performed various numerical experiments and we summarise the results here:

**Experiment (i)** Structure: Annealed TPL70 network and homogeneous network with same mean degree. Disease parameters: Identical recovery rate, intrinsic generation interval (*γ* = 1) and ℛ_0_. Result: Heterogeneity reduces the contraction in the forward generation interval. However, this could possibly be explained by the lower peak (hence lesser competition) in the heterogeneity (See supplementary results).

**Experiment (ii)** Structure: Annealed TPL70 network and homogeneous network with same mean degree.

Disease parameters: Identical recovery rate, intrinsic generation interval (*γ* = 1). The transmission rate is selected such that the two epidemics have the same level of peak incidence of infections. Result: Heterogeneity increases the contraction in the forward generation interval, despite the same maximum incidence of infections. This can be explained by noting that the peak of *π*_*i*_(*t*) is higher than *i*(*t*). Furthermore, the backward generation intervals also differ significantly (See supplementary results).

**Experiment (iii)** Structure: Annealed TPL70 network Method: Compare distribution of realised intervals assuming homogeneity [17] (i.e. using *i*(*t*) and *S*(*t*)) and not assuming homogeneity (i.e. using *π*_*i*_(*t*) and *π*_*S*_(*t*))

Result: Using the net incidence of infections underestimates the contraction in forward generation interval. In contrast, the backward generation intervals are relatively unchanged (See supplementary results).

### 3.4 Implications for estimation of reproduction number

In this section, we will explore how errors could arise if disease parameters are inferred using existing methods to relate the observed generation interval and growth rate to the basic reproduction number.

If the intrinsic generation intervals and the exponential growth rate are known, the basic reproduction number can be estimated using the Wallinga-Lipsitch equation [15]. The growth rate can be estimated from incidence at the start of the epidemic. However, the intrinsic generation intervals are not observed. Instead we observe the realised generation intervals (forward or backward) which are influenced by the dynamics of the outbreak and the contact structure of the population.

So to fit model parameters and infer the basic reproduction number from the growth rate and the observed generation intervals, it is crucial to recognise the distinction between the realised generation intervals and the intrinsic generation intervals. In principle, the exact expressions for realised generation interval distributions can be used to infer the intrinsic generation interval distribution, which can be used to then estimate ℛ_0_ [17, 5, 14, 6]. However, the Champredon and Dushoff’s equation and further developments relies on the homogenous mixing assumption.

Our results from previous sectionss show that for a heterogeneous population, the conclusions will be incorrect if simple population aggregates such as prevalence of susceptibles or incidence of infections are used. We expect that the joint use of simple aggregate observations of an outbreak and realised generation intervals could bias the estimation of the basic reproduction number and the intrinsic generation intervals. To demonstrate this bias, we now look at an example case of outbreak analysis and two scenarios with different amounts of information available to the disease modeller.

**Example** Let us consider a pathogen causing an epidemic in different disconnected populations. There is a population in which the authorities have the capacity and resources to document the incidence of cases and also perform backward contact tracing (let us call it Population 1). There is another population where we do not have the capacity to perform contact tracing, but we can document case incidence (call it Population 2). Both populations are well described by a quenched and heterogeneous contact network. This is a simplification, as any real-world network will have a mixture of short and long time scale contacts. First, we describe the quantities of interest in the ground reality from the correct model. Then, we consider cases where an infectious disease modeller has some information about the contact structure and see how their modelling assumptions can affect model inference in both populations.

Using the ground reality model, we can write the basic reproduction numbers in terms of the observable quantities:

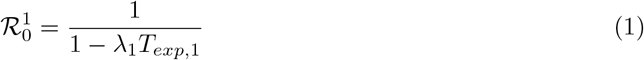

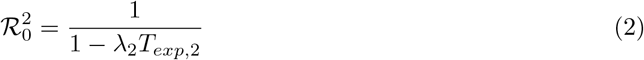

where the *λ* is the exponential growth rate estimated from incidence and *T*_*exp*_ is the mean backward generation interval during the exponential growth phase, estimated from the contact tracing data. We can also write the average intrinsic generation time as a function of the same quantities. However, the expression depends also on the first and second moments of the degree distribution. See detailed methods for a detailed calculation.

**Case A:** The modeller does not know anything about the contact structure and assumes a well-mixed homogeneous model. The model has essentially two parameters, the transmission rate *β* and the recovery rate *γ* (which also specifies the intrinsic generation intervals). Using this model, the estimators for population 1 are:

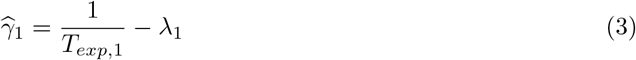

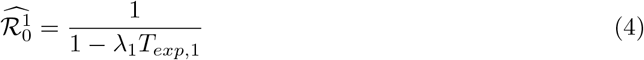

Comparing these estimates to the ground reality tells us that the basic reproduction number is unbiased but 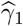 is not as it does not properly account for the contact network properties – first and second moment of the degree distribution.

For population 2, nothing is known about the backward generation intervals, but the modeller has an estimate for the intrinsic generation intervals in population 1. Since by definition, intrinsic intervals would not vary among populations, the modeller might see it fit to treat 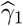 as an estimator for *γ*_2_. This leads to an estimate of the reproduction number

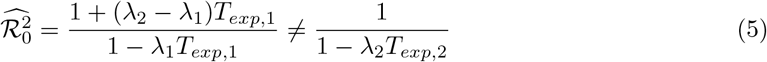

which is biased. We find that the bias in the estimator for population 2 is greater when the contact structure is different among the two populations. When the contact structure is identical, the estimator is unbiased when the growth rates are equal.

**Case B:** The modeller knows that the contacts are quenched and also has correct information of the average number of contacts ⟨*K*_1_⟩ and ⟨*K*_2_⟩ respectively. In the absence of more detailed information, they assume a homogeneous quenched network with degrees ⟨*k*_1_ = *K*_1_⟩ and ⟨*k*_2_ = *K*_2_⟩, respectively. The model has two other parameters, the transmission rate *β* and the recovery rate *γ* (which also specifies the intrinsic generation intervals). Using this model, the estimators for population 1 are:

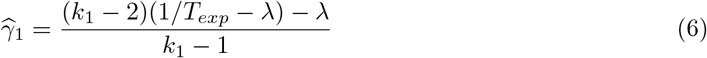

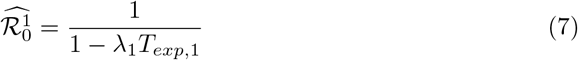

Comparison with the ground reality model shows that 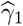 is biased, as calculation does not correctly account for the second moment of the degree distribution, while 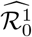 is not. As in the previous case, the modeller might see fit to treat 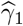 as an estimator for *γ*_2_. This leads to an estimate of the reproduction number for population 2

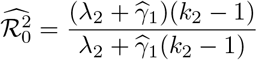

This estimator is biased, albeit behaves very differently than the biased estimator in Case A.

In summary, we find that if the incidence and backward contact tracing is available in a population, then even incorrect assumptions about the contact structure can yield an unbiased estimator for the basic reproduction number, even though the estimator for the intrinsic generation interval is biased. Using this estimated intrinsic generation interval in order to estimate the basic reproduction number in another population leads to biased estimates. See 3 for an illustration of the bias in 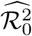.

While case A is realistic, case B is hypothetical since more information – i.e. the average number of contacts – is available and used in the analysis than what normally happens in reality. Paradoxically, case B presents a more severe bias. For Population 1, a truncated power law degree distribution is used with an exponent of −2 and maximum degree of 70. Population 2 has a similar structure but with a maximum degree of 30. The growth rate in Population 1 is *λ*_1_ = 0.5 and recovery rate is *γ* = 1.

## 4 Discussion

In this article, we have generalised Champredon and Dushoff’s theory [17] for realised generation intervals in a homogeneous and well-mixed population to heterogeneous networks, using the EBCM framework. This framework allows incorporating degree heterogeneity in a tree-like network. In structured populations, realised generation intervals have been investigated using network and agent-based simulations [19, 4]. These methods benefit from their ability to be modified quite easily – adding more coarse-grained structure or even including forms of structure beyond degree heterogeneity. However, they are slow and managing the noise reasonably would require a large number of simulations or a large population. In addition, they do not provide analytical insights. Some analytical considerations were provided by a few works [20, 11]. Our exact expressions, validated using simulations, present an advance in our ability to predict generation intervals in simple heterogeneous contact structures and sets up a foundation to obtain exact expressions for more complex heterogeneous networks.

Generation intervals are useful in estimating the basic reproduction number of an epidemic. Champredon and Dushoff showed how fitting the mean intrinsic generation interval to the mean of the realised generation intervals (backward) is incorrect [17]. They anticipate that heterogeneity may affect their results. We confirm this, determine the manner by which it affects the results and more importantly, through what mechanism. We found that the realised intervals depend not on simple aggregate measures like net prevalence of susceptibles or net incidence of infection, but on more involved measures such as the prevalence of half-edges in the network which have a susceptible or a newly infected node attached to them. Measurement of such quantities requires higher resolution observation of the population, at the scale of groups which are homogeneous in number of contacts. We also use a series of hypothetical experiments to show that the estimates of intrinsic intervals (inferred using realised generation intervals) are sensitive to model assumptions. Despite this sensitivity, the estimates of ℛ_0_ can be relied upon as long as the estimated intrinsic generation intervals are used in the same setting where they were estimated. If the estimated intrinsic intervals are used in a different setting (say an epidemic caused by the same pathogen in a different setting where the contact/network structure differs) then the estimates of ℛ_0_ will be biased. This practice of applying generation interval estimates or related parameter estimates from from one setting to an epidemic of the same pathogen in another setting is common [22, 23, 24, 25, 26, 27].

The forward reproduction number that was derived here is an effective reproduction number. Effective reproduction numbers are of two types: instantaneous or cohort-based (case reproduction number). The instantaneous reproduction number measures the transmission happening at certain time while the case reproduction number measures the transmission that is caused by a cohort of infectors who became infected at a certain time [28]. The forward reproduction number, ℛ_*f*_, is equivalent to the case reproduction number [28, 29].

A key assumption throughout this work is that the recovery from infection is a Poisson process (the intrinsic generation interval distribution is exponential). While real-world diseases do not follow this assumption, the expressions for forward and backward generation intervals that we derived are fairly general. Real-world networks have many kinds of heterogeneities beyond the kind we have investigated here. They have clustering (triads that form a triangle), multiple layers and temporal variation. Here, we have only studied degree heterogeneity in two extreme cases of temporal variation – quenched (static partners) and annealed (fleeting contact with changing partners). In principle, a linear combination of the distributions derived here may be used to model contact networks with a more realistic temporal behaviour.

## Supporting information

Supplementary methods and results

## Data Availability

All software used to generate the simulations are available online at https://github.com/praty-k/gi-networks

https://github.com/praty-k/gi-networks

## Data and Software

The software used in this study is available at https://github.com/praty-k/gi-networks

## Acknowledgements

We thank Andrew Black and Eben Kenah for helpful conversations. This research was supported by use of the Nectar Research Cloud, a collaborative Australian research platform supported by the NCRIS-funded Australian Research Data Commons (ARDC). P.K.K. was supported by the La Trobe Graduate Research Scholarship and Full-Fee Research Scholarship and the Ed Smith Applied Mathematics Fellowship. C.P. was supported by the Cariparo Foundation through the program Starting Package and the Department of Molecular Medicine through the program SID from BIRD funding.

