## Supplementary methods and results for "Network structure induced bias in estimates of intrinsic generation times"

### 1 Supplementary Methods

We use the framework of Edge Based Compartmental Models to derive exact expressions for the generation interval distribution [2]. We assume that the infectiousness, $\beta$, is constant, and that infectious individuals recover at a constant rate $\gamma$. Unless otherwise stated, the heterogeneous network refers to a configuration model network with a truncated power law degree distribution which has an exponent of $-2$, and integer degrees between 1 and 70. The mean degree of nodes in such a network is $\sim 3$ and the mean of square of degrees is $\sim 43$. These properties are used in calculating the basic reproduction number, growth rate etc. [2]

#### 1.1 Annealed Network

The average probability of contacting an individual in state $X$ who has $k$ stubs (or is of degree $k$) is proportional to the product of probability of being in state $X$, of having $k$ stubs, and the number of stubs $k$ (because if a node has more stubs, then a random node is more likely to connect to it):

$$P(X,k) \sim X_k k, \text{and } \pi_X = \frac{\sum_k X_k k}{\sum_k N_k k}, \text{Thus, } \pi_S(t) = \frac{\sum_k S_k k}{\sum_k N_k k} = \frac{\sum_k N_k \theta^k k}{\sum_k N_k k} = \frac{\theta\psi'(\theta)}{\psi'(1)}$$

where $\theta$ is the probability that the neighbour of a randomly selected node has not transmitted to it yet. So, the number of susceptibles of degree $k$ is equal to the number of individuals of degree $k$, times the probability that none of its $k$ neighbours has transmitted to it. The probability with which a randomly selected individual would come in contact with the stub of a susceptible node is

$\pi_S$, with the stub of an infected node is $\pi_I$, with the stub of a recovered node is $\pi_R$ [2]. These three probabilities would sum up to unity.

Using the EBCM framework, we define new a quantity, $\pi_i(t)$, which is the probability of contacting the stub of a newly infected node at time $t$. This can also be described as the average incidence of infected stubs.

The depletion rate of susceptibles of degree $k$ is given by:

$$\frac{dS_k}{dt} = -\beta k S_k(t)\pi_I(t)$$

This is also negative of the rate at which new infections are being created. At time $t$, the creation rate (i.e., the incidence) of new infections of degree $k$ is

$$i_k(t) = \beta k S_k \pi_I(t)$$

The rate of contacting an infectious individual who was infected at time $t$ and has $k$ edges will be $ki_k(t)$. And the rate of contacting an individual infected at $t$ will be the average $\frac{\sum_k k i_k}{\langle k \rangle}$. The incidence of infections of degree $k$ is $-\frac{dS_k}{dt}$. Therefore, from equation (), the average incidence of stubs can be defined as

$$\pi_i = \beta\pi_I \frac{\sum_k k^2 S_k}{\langle k \rangle} = \beta\pi_I \frac{\sum_k k^2 N_k \theta^k}{\langle k \rangle} = \beta\pi_I \frac{\theta^2\psi\text{''}(\theta) + \theta\psi'(\theta)}{\psi'(1)}$$

The incidence of infected stubs can also be obtained by considering the fact that the incidence of infected stubs is the same as the rate at which $\pi_S$ decreases (since nodes, and hence their stubs, can only transition from state-$S$ to state-$I$). Using the EBCM framework, an expression for $-\frac{d\pi_S}{dt}$ can be derived, which is equivalent to the expression for $\pi_i$ derived above [2].

**Forward Generation Interval Distribution** In the homogeneous case, the number of new cases at time $t+\tau$ caused by an average person infected at time $t$ is proportional to [1]

$$i_t(t+\tau) \sim g(\tau)S(t+\tau)i(t)$$

For a configuration model, the number of new cases at time $t+\tau$ with degree $k$ caused by people with degree $l$ infected at time $t$ is proportional to

$$g(\tau)S_k(t+\tau)ki_l(t)l$$

So the incidence at time $t+\tau$ caused by the average person infected at time $t$ is

$$i_t(t+\tau) \sim g(\tau)\sum_k kS_k(t+\tau)\sum_l li_l(t) = g(\tau)\pi_S(t+\tau)\sum_l li_l(t)$$

The distribution of forward generation intervals ($\tau$) at time $t$ is obtained by normalising the above derived incidence:

$$f_A(t,\tau) = \frac{g(\tau)\pi_S(t+\tau)}{\int_0^\infty dx\ g(x)\pi_S(t+x)}$$

**Backward Generation Interval Distribution** In the homogeneous case, the number of new cases at time $t$ caused by an average person infected at time $t-\tau$ (i.e. $\tau$ units ago) is proportional to [1]

$$i_{t-\tau}(t) \sim g(\tau)S(t)i(t-\tau)$$

For a configuration model, the number of new cases at time $t$ with degree $k$ caused by people with degree $l$ infected at time $t-\tau$ is proportional to

$$g(\tau)S_k(t)ki_l(t-\tau)l$$

So the incidence at time $t$ caused by the average person infected at time $t-\tau$ is

$$i_{t-\tau}(t) \sim g(\tau)\sum_k kS_k(t)\sum_l li_l(t-\tau) \sim g(\tau)\pi_S(t)\pi_i(t-\tau)$$

Normalising this incidence derived above will yield the backward generation interval distribution at time $t$:

$$b_A(t,\tau) = \frac{g(\tau)\pi_i(t-\tau)}{\int_0^\infty dx\ g(x)\pi_i(t-x)}$$

### 1.2 Quenched Network

A quenched network differs from an annealed/well-mixed/mean-field network in that the partnerships are static in time. In a well-mixed network, a node does not compete with itself to infect a certain node since the edges rewire constantly. In the quenched case, if an infected node transmits to a (susceptible) neighbour twice, then only the first transmission counts as an infection event. The second transmission does not lead to an infection. Thus, the generation intervals are shorter compared to the well-mixed case even when there are no other competing infectors. When calculating the realised (forward and backward) generation intervals, we need to use an effective intrinsic generation interval which is equivalent to the distribution of first transmissions between a pair of infected and susceptible partners.

If the intrinsic generation intervals are given by

$$g(t) = \gamma e^{-\gamma t}$$

then the effective intrinsic generation interval is

$$\tilde{g}(t) = (\beta+\gamma)e^{-(\beta+\gamma)t}$$

This can be derived by considering the first transmission event between a pair of connected nodes. The rate at which the first transmission event occurs at time $t$ is the product of the probabilities of the infector being infectious ($e^{-\gamma t}$), the infector not yet transmitting ($e^{-\beta t}$), the rate of transmission $\beta$. Normalising this product gives the rate at which first transmissions occur.

As the above derived equation shows, the first transmission event is a poisson process with rate $\beta+\gamma$. Instead of $\tilde{g}(t)$, what we will be using from here on is the probability that the first transmission has not occurred yet:

$$\tilde{G}(t) = e^{-(\beta+\gamma)t}$$

**Forward Reproduction Number** For the quenched network, we will derive a forward reproduction number using the renewal equation. Fundamentally, the renewal equations relates the incidence at present to the incidence during the time that has passed. If the present time is $t$, then

$$\begin{gathered}\text{Incidence }(t) = \int_0^t \sum_k \text{Incidence of degree } k \text{ at time } (t-\tau)\\ \times\text{Rate of new infections produced by a single infector of}\\ \text{degree } k \text{ who was infected } \tau \text{ units ago}\end{gathered}$$

Let us calculate the second term of the product. The number of new infections created at time $t+\tau$, by a node $u$, of degree $k$, infected at time $t$ ($\tau$ units ago) can be computed by:

$$\begin{gathered}P(u \text{ is infectious at } t+\tau \text{ given it became infected at } t)\\ \times P(u \text{ didn't transmit until } t+\tau \text{given it became infected at } t)\\ \times(\text{No. of neighbours it could infect})\\ \times(\text{Rate at which } u \text{ transmits to a neighbour})\\ \times P(\text{a neighbour } v \text{ is susceptible at } t+\tau \text{ given } u \text{ was susceptible at time } t)\\ = e^{-(\beta+\gamma)\tau} \times (k-1) \times \beta \times P(v_{t+\tau} = S \mid u_t = S)\end{gathered}$$

$$P(v_{t+\tau} = S \mid u_t = S) = \tfrac{P(v_{t+\tau}=S \cap u_t=S)}{P(u_t=S)}$$

The denominator, $P(u_t = S)$ is $\theta^k(t)$. The numerator can be written as

$$\begin{gathered}P(v_{t+\tau} = S \cap u_t = S) = P(v_{t+\tau} = S \cap v \nrightarrow u \text{ by time } t)\\ \times P(k-1 \text{ neighbours } \nrightarrow u \text{ by time } t)\\ = \phi_S(t+\tau)\theta^{k-1}(t)\end{gathered}$$

where $v \nrightarrow u$ denotes $v$ did not transmit to $u$. Therefore,

$$P(v_{t+\tau} = S \mid u_t = S) = \frac{\phi_S(t+\tau)}{\theta(t)}$$

The number of new infections generated at time $t+\tau$, by a node of degree $k$ who was infected at time $t$, is:

$$\beta(k-1)e^{-(\beta+\gamma)\tau}\frac{\phi_S(t+\tau)}{\theta(t)}$$

Next, let us try to find the incidence at time $t$ in a degree class $k$. The probability of getting infected at time $t$ given that a node $u$ has degree $k$ is

$$\begin{gathered}P(u \text{ is susceptible at } t)\\ \times \text{ Rate at which an infected neighbour transmits}\\ \times \text{ Number of neighbours who could transmit to } u\\ \times\text{Probability that a neighbour } v \text{ is infected and has not transmitted to } u\\ \text{given that } u \text{ is susceptible}\\ = \theta^k(t) \times \beta \times k \times P(v_t = I \ \cap \ v \nrightarrow u \mid u_t = S)\end{gathered}$$

$$P(v_t = I \ \cap \ v \nrightarrow u \mid u_t = S) = \frac{P(v_t=I \ \cap \ v \nrightarrow u \ \cap \ u_t=S)}{P(u_t=S)}$$
$$= \frac{P(v_t=I \ \cap \ v \nrightarrow u) \times P(k-1 \text{ neighbours } \nrightarrow u \text{ by time } t)}{P(u_t=S)}$$
$$= \frac{\phi_I(t)}{\theta(t)}$$

Therefore, the probability of getting infected at time $t$ given that the node has degree $k$ is

$$\beta k \phi_I(t) \theta^{k-1}(t)$$

Total incidence at time $t$ is therefore

$$N\beta\phi_I(t) \sum_k k N_k \theta^{k-1}(t) = N\beta\phi_I(t)\psi'(\theta(t))$$

where $N$ is the population size.

Now, let us find the total number of new infections at time $t+\tau$ created by individuals who got infected at time $t$ ($\tau$ units ago). We make use of the two quantities dervied above: incidence of degree $k$ at time $t$ and number of infections created at time $t+\tau$ by a single infector of degree $k$ who was infected at time $t$.

$$N \sum_k \beta k N_k \theta^{k-1}(t)\phi_I(t) \times \beta(k-1)e^{-(\beta+\gamma)\tau} \frac{\phi_S(t+\tau)}{\theta(t)}$$
$$= N\beta^2\phi_I(t)\phi_S(t+\tau)e^{-(\beta+\gamma)\tau} \sum_k k(k-1)N_k\theta^{k-2}(t)$$
$$= N\beta^2\phi_I(t)\phi_S(t+\tau)e^{-(\beta+\gamma)\tau}\psi''(\theta(t))$$

Dividing this by the total incidence at time $t$ and integrating over $\tau$ gives the expected number of infections created over an infectious period by the average individual who was infected at time $t$.

$$\frac{\beta\psi''(\theta(t))}{\psi'(\theta(t))} \int_0^\infty d\tau \phi_S(t+\tau)e^{-(\beta+\gamma)\tau}$$

We call this quantity the forward reproduction number, in analogy to the forward generation interval. Note that the term in the product, $\frac{\psi''(\theta(t))}{\psi'(\theta(t))}$ depends only on $\theta$ and the degree distribution. It can be interpreted as the expected degree of a node that got infected at time $t$.

**Forward Generation Interval Distribution** The forward generation interval distribution at time $t$ can be computed using the rates (as a function of time $\tau$) at which new infections are produced by the infections which happened at $t$. This rate is proportional to the probability that a partner of a randomly selected node is susceptible, $\phi_S$, and the probability that the selected node is still infectious in the future, given it becomes infected at present time, and the probability that it hasn't transmitted to this partner before. The distribution is obtained by normalising these rates against $\tau$.

In a quenched network, the rate at which new infections are caused at time $t+\tau$, by infections which were generated at time $t$

$$i(t+\tau) \sim \tilde{G}(\tau)\phi_S(t+\tau)$$

because $\phi_S(t+\tau)$ is the probability that an infected neighbour of a susceptible node transmits and $\tilde{G}(\tau) = e^{-(\beta+\gamma)\tau}$ is the probability that a node infected $\tau$ time units ago would emit its first transmission. Therefore, the forward generation interval distribution will be given by

$$f_Q(t,\tau) = \frac{\tilde{G}(\tau)\phi_S(t+\tau)}{\int_0^\infty dx\ \tilde{G}(x)\phi_S(t+x)}$$

**Backward Generation Interval Distribution** The rate at which new infections are caused at time $t$ due to infections created $\tau$ units ago (at time $t-\tau$) is

$$i_{t-\tau}(t) \sim \tilde{G}(\tau)\phi_i(t-\tau)$$

because $\phi_i(t-\tau)$ is the rate at which a susceptible neighbour of a node gets infected at time $t-\tau$ and enters the $\phi_I$ state (i.e. infected neighbour that has not yet transmitted to our node of interest) and $g(\tau)$ is the probability density that a node infected $\tau$ units ago would emit its first transmission. Therefore, the backward generation interval can be obtained by normalising this rate

$$b_Q(t,\tau) = \frac{\tilde{G}(\tau)\phi_i(t-\tau)}{\int_0^\infty dx\ \tilde{G}(x)\phi_i(t-x)}$$

#### 1.2.1 Estimation of reproduction number

**Ground Reality:** In Population I, the parameters are $\beta_1$, $\gamma$ and $K_1$, which are the transmission rate, recovery rate (also specifies the intrinsic generation interval) and the random variable for the number of contacts. The growth rate is $\lambda_1$.

$$\lambda_1 = \beta_1 \frac{\langle K_1^2 \rangle}{\langle K_1 \rangle} - 2\beta_1 - \gamma \tag{1}$$

$$\mathcal{R}_0^1 = 1 + \frac{\lambda_1}{\beta_1 + \gamma} \tag{2}$$

Similarly, for Population 2,

$$\lambda_2 = \beta_2 \frac{\langle K_2^2 \rangle}{\langle K_2 \rangle} - 2\beta_2 - \gamma \tag{3}$$

$$\mathcal{R}_0^2 = 1 + \frac{\lambda_2}{\beta_2 + \gamma} \tag{4}$$

The recovery rate is identical for both populations because it defines the intrinsic generation intervals, which by definition, are not affected by changes in contact structure. Using the expression for backward generation intervals, the mean backward generation interval can be calculated when the epidemic is growing exponentially

$$T_{exp,1} = \frac{1}{\lambda_1 + \beta_1 + \gamma} \tag{5}$$

$$T_{exp,2} = \frac{1}{\lambda_2 + \beta_2 + \gamma} \tag{6}$$

The quantities of interest, which are the basic reproduction number and the intrinsic generation interval, can now be expressed in terms of the observable variables

$$\mathcal{R}_0^1 = \frac{1}{1 - \lambda_1 T_{exp,1}} \tag{7}$$

$$\mathcal{R}_0^2 = \frac{1}{1 - \lambda_2 T_{exp,2}} \tag{8}$$

$$\gamma = \frac{\left(\frac{\langle K_1^2 \rangle}{\langle K_1 \rangle} - 2\right)\left(\frac{1}{T_{exp,1}} - \lambda_1\right) - \lambda_1}{\frac{\langle K_1^2 \rangle}{\langle K_1 \rangle} - 1} \tag{9}$$

$$= \frac{\left(\frac{\langle K_2^2 \rangle}{\langle K_2 \rangle} - 2\right)\left(\frac{1}{T_{exp,2}} - \lambda_2\right) - \lambda_2}{\frac{\langle K_2^2 \rangle}{\langle K_2 \rangle} - 1} \tag{10}$$

**Case A:** The modeller does not know anything about the contact structure and decides to use the simplest model available, the homogeneous and well-mixed SIR scheme to model these epidemics. This model has two parameters, the contact rate $\beta$ and the recovery rate $\gamma$. The recovery rate also specifies the intrinsic generation interval distribution. The exponential growth rate in this model is $\beta - \gamma$ and this rate can be obtained from the incidence.

$$\lambda = \beta_1 - \gamma_1$$

Using the expression for backward generation intervals, under the assumption that incidence grows exponentially with rate $\lambda$, the mean backward generation interval, $T_{exp}$ is

$$T_{exp} = \frac{1}{\lambda + \gamma_1}$$

Since, we have access to backward contact tracing, $T_{exp,1}$ is obtained from the data. Using these two equations in conjunction, the parameters $\beta$ and $\gamma$ can be estimated, along with the basic reproduction number.

$$\widehat{\beta}_1 = \frac{1}{T_{exp,1}} \tag{11}$$

$$\widehat{\gamma}_1 = \frac{1}{T_{exp,1}} - \lambda_1 = \beta_1 + \gamma \tag{12}$$

$$\widehat{\mathcal{R}_0^1} = 1 + \frac{\lambda_1}{\widehat{\gamma}_1} = \frac{1}{1 - \lambda_1 T_{exp,1}} \tag{13}$$

The estimated distribution of intrinsic generation intervals is incorrect, yet the estimate for the basic reproduction number is correct.

By definition, the intrinsic generation interval distribution depends solely on how the infectiousness of an infected person changes with time. It does not depend on contact structure or the availability of susceptible contacts. Therefore, the modeller might see it fit to use the estimated intrinsic interval distribution from Population 1 to estimate the basic reproduction number in Population 2 as they do not have contact tracing capacity there.

$$\widehat{\mathcal{R}_0^2} = 1 + \frac{\lambda_2}{\widehat{\gamma}_1} \tag{14}$$

$$\widehat{\mathcal{R}_0^2} = \frac{1 + (\lambda_2 - \lambda_1)T_{exp,1}}{1 - \lambda_1 T_{exp,1}} \neq \frac{1}{1 - \lambda_2 T_{exp,2}} \tag{15}$$

This estimate of the basic reproduction number is different from the true reproduction number unless the growth rates in the populations are equal ($\lambda_2 = \lambda_1$), for which there is no a priori justification.

This example can also be framed in a different manner. Say there are two populations which are similar in their contact structure, but differ in their implementation of interventions.

**Case B:** The modeller knows that the population is well described by a quenched network and knows the average degrees of the two populations, so they assume a quenched homogeneous model. This model has three parameters, the contact rate $\beta$, the recovery rate $\gamma$ and the degree of nodes, $k$ (which is known). The exponential growth rate in this model is $\beta(k-2) - \gamma$ and this rate can be obtained from the incidence.

$$\lambda_1 = \beta_1(k_1 - 2) - \gamma_1$$

Using the expression for backward generation intervals, under the assumption that incidence grows exponentially with rate $\lambda$, the mean backward generation interval, $T_{exp,1}$ is

$$T_{exp,1} = \frac{1}{\lambda_1 + \gamma_1 + \beta_1}$$

Since, we have access to backward contact tracing, $T_{exp,1}$ is obtained from the data. Using these two equations in conjunction, $\gamma_1$ can be estimated, along with the basic reproduction number.

$$\widehat{\gamma}_1 = \frac{(k_1 - 2)(1/T_{exp} - \lambda) - \lambda}{k_1 - 1} \tag{16}$$

$$\widehat{\mathcal{R}_0^1} = 1 + \frac{\lambda_1}{\widehat{\gamma}_1 + \widehat{\beta}_1} = \frac{1}{1 - \lambda_1 T_{exp,1}} \tag{17}$$

The estimated distribution of intrinsic generation intervals is incorrect, but the estimate for the basic reproduction number is correct. The intrinsic generation interval can be estimated properly only when the degree of the homogeneous model matches the ratio of mean square degree and mean degree from the actual contact network.

As in the previous case, the estimated intrinsic generation intervals would be used to understand the dynamics in Population 2, which would lead to

$$\widehat{\mathcal{R}_0^2} = 1 + \frac{\lambda_2}{\widehat{\beta}_2 + \widehat{\gamma}_1} = \frac{(\lambda_2 + \widehat{\gamma}_1)(k_2 - 1)}{\lambda_2 + \widehat{\gamma}_1(k_2 - 1)} \tag{18}$$

This estimate of the basic reproduction number is different from the actual basic reproduction number in Population 2.

In this example, we do not fit the full time series of mean backward intervals to the models, instead we focus on a section of the time series where the behaviour is analytically tractable. We would guess that fitting the full time series would present more challenges and bias the estimators further.

Insert plot of $\widehat{\mathcal{R}_{0,II}}$ and $\mathcal{R}_{0,II}$ with $\beta_2$ on axis to show the estimation error.

### 2 Supplementary Results

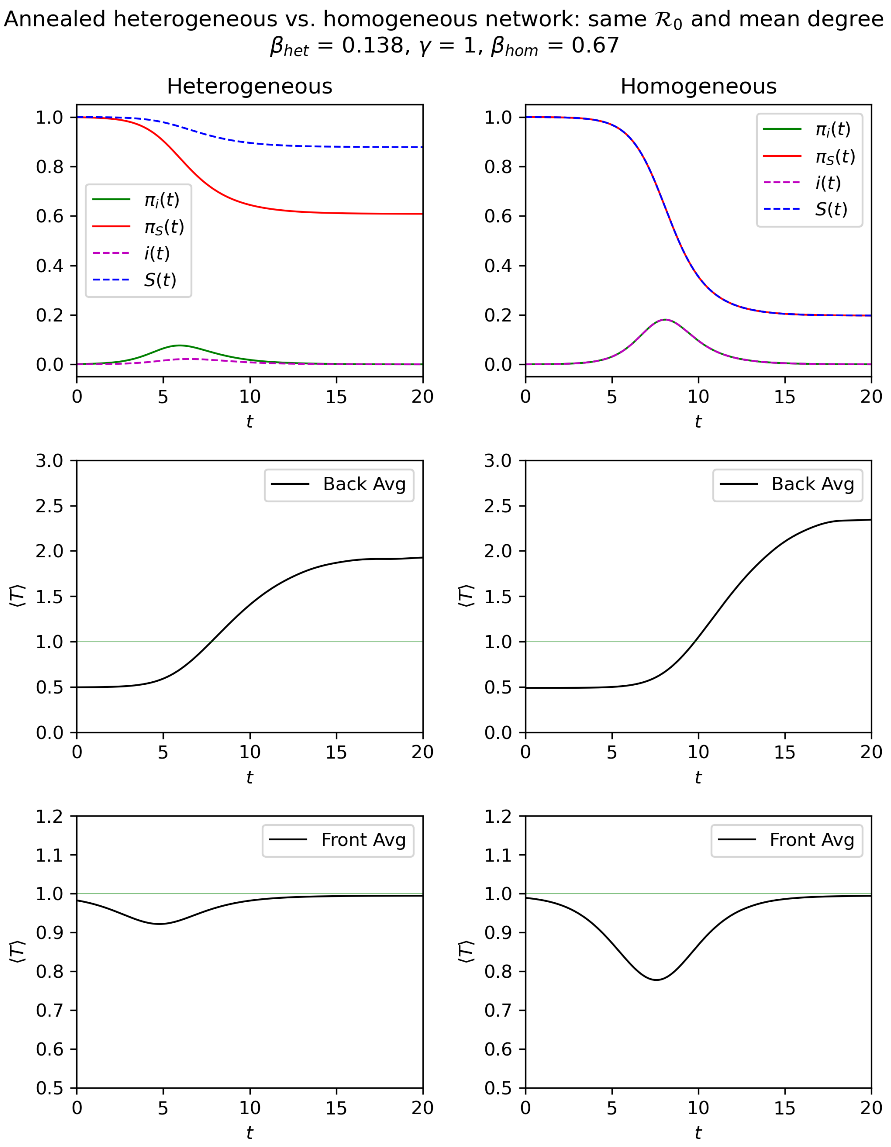


**Figure S1** *Epidemics and their realised generation intervals for two networks: a heterogeneous annealed network and a homogeneous annealed network. Parameters are selected such that basic reproduction number and intrinsic generation interval distribution are the same for both epidemics. Yet, the realised intervals look different. The heterogeneity clearly decreases the amount of con-*

*traction in the mean forward generation interval. This can be explained by looking at the incidence of stubs,* $\pi_i(t)$*. In the homogeneous case, the peak is higher than the heterogeneous epidemic. More competition among infectors shortens the generation time more. The starting time,* $t = 0$*, is selected such that the prevalence is* $0.01\%$*, and it can be seen that in the heterogeneous case, the mean generation interval is shorter at the start. This can be explained by the faster fall in the prevalence of susceptible stubs in the population,* $\pi_S(t)$*. The mean backward generation interval is quite similar at early times, and deviates at a later time. This can be explained by the same early exponential growth rate and different rates of exponential decline*

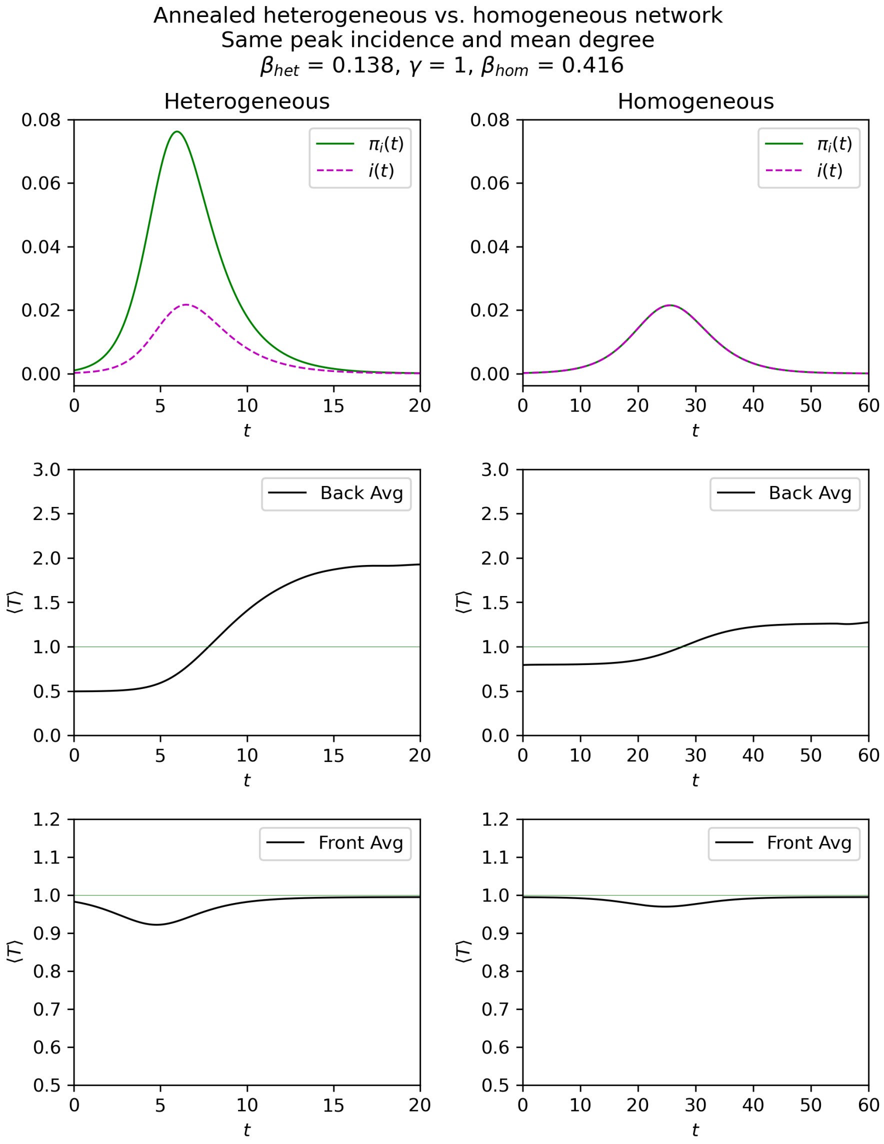


**Figure S2** *Epidemics and their realised generation intervals for two networks: a heterogeneous annealed network and the other a homogeneous annealed network, with identical mean degree. Parameters are selected such that the peak incidence and intrinsic generation interval distribution are the same for both epidemics. Yet, the realised intervals look very different.*

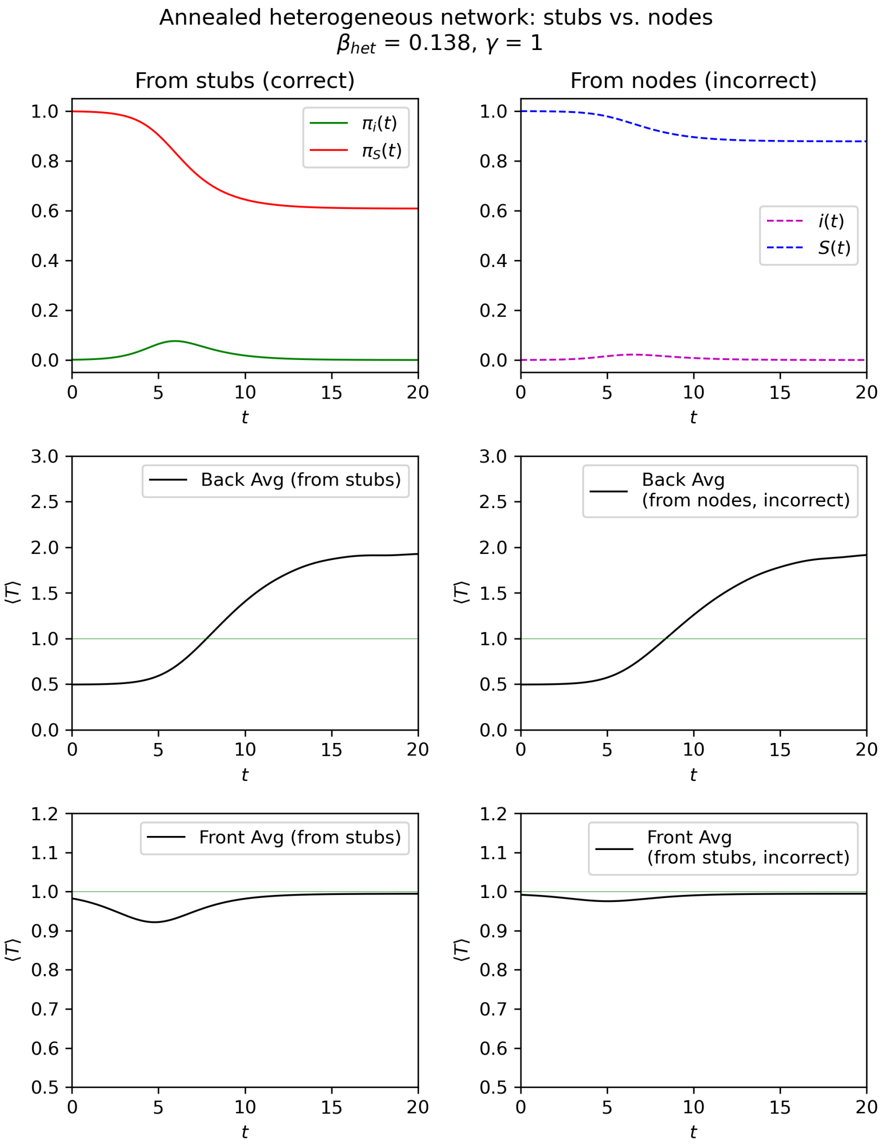


**Figure S3** *An epidemic and its mean realised generation intervals on an annealed network with truncated power law degree distribution. The realised generation intervals calculated using the equations from [1] and the equations we derived are markedly different. The backward mean generation intervals are identical at the start of the epidemic. The homogeneous assumption underestimates the contraction in forward generation intervals.*
